# Scan length as a major driver of CT radiation dose: a diagnostic reference level audit from Kosovo

**DOI:** 10.64898/2026.05.12.26353024

**Authors:** Granit Rudi, Fator Vula, Ardian Bicaku, Kreshnike Dedushi Hoti, Ilir Ahmetgjekaj

**Author notes:** **Corresponding author:** Fator Vula, MD, Clinic of Radiology, University Clinical Centre of Kosovo, Pristina, 10000, Kosovo.

## Abstract

Computed tomography is the largest contributor to population radiation dose from medical imaging, yet no diagnostic reference levels (DRLs) have been published from Kosovo or the Western Balkans. This retrospective audit analyzed all CT examinations performed on a 128-slice scanner at the University Clinical Centre of Kosovo between January and March 2026. After exclusions, 1,535 acquisitions from 1,092 patients across nine examination categories were analyzed. Local DRLs were defined as the 75th percentile and compared against German (BfS 2022) and Turkish (Kahraman et al., 2024) reference values. Head CT (n = 590) demonstrated CTDIvol 4.7% below the BfS DRL yet scan length 98.5% above the orientation value (median 25.8 vs 13 cm). Abdomen-pelvis CTDIvol matched the BfS reference while scan length exceeded it by 28%. Coronary CTA showed CTDIvol +377%, consistent with retrospective ECG gating. Excess scan length, not CTDIvol, is the major driver of elevated dose at this institution. The identified excesses are correctable through technologist landmarking training, protocol review, and enabling iterative reconstruction.

## Introduction

Computed tomography is the single largest contributor to population radiation dose from medical imaging. Although CT accounts for approximately 10% of all diagnostic imaging procedures worldwide, it contributes an estimated 63% of the collective effective dose from medical exposures. (UNSCEAR, 2022; Garba et al., 2021) The optimization of CT radiation dose is therefore a central objective of radiological protection, operationalized through the diagnostic reference level (DRL) framework established by the International Commission on Radiological Protection. (Vañó et al., 2017; ICRP, 2007) Local DRLs are recommended as a starting point where national values do not exist. (Damilakis et al., 2023)

DRLs are set at the 75th percentile of the dose distribution for a given examination type across a sample of facilities, providing a benchmark against which individual institutions can compare their practice. (Vañó et al., 2017) They are not dose limits but investigation triggers: facilities consistently exceeding the DRL are expected to review their protocols and identify opportunities for optimization. Scan length has previously been reported as a complementary dose metric in multi-center benchmarking studies. (Pyfferoen et al., 2017) This iterative audit-and-feedback process has been demonstrated to reduce both the magnitude and variability of CT doses in countries with established DRL programs. (Vañó et al., 2017) The European Basic Safety Standards Directive requires all member states to establish, regularly review, and use DRLs for common diagnostic procedures. (Council Directive 2013/59/Euratom, 2014)

Despite this regulatory framework, substantial gaps remain. No CT diagnostic reference levels — national or local — have been established in Kosovo. This absence extends to the broader Western Balkans region, where no published CT DRL data exist for Albania, North Macedonia, Montenegro, Bosnia and Herzegovina or Serbia. Kosovo therefore lacks the baseline data necessary to identify whether its CT practice requires optimization, and if so, where.

A further limitation of most existing DRL frameworks is their reliance on a single dose metric. Traditionally, DRLs have been expressed as CTDIvol or dose-length product (DLP). However, DLP alone does not distinguish between high beam intensity over a short scan and low beam intensity over an excessively long scan — two scenarios with different optimization targets. Recognizing this, the German Federal Office for Radiation Protection (BfS) updated its national DRL framework in 2022 to report CTDIvol as the formal DRL and scan length as a separate orientation parameter, replacing DLP entirely. (BfS, 2022) The BfS explicitly noted that this was the first time scan range and scan length were included as orientation values for CT examinations. (BfS, 2022) This shift acknowledges that scan coverage is a critical but under-monitored determinant of patient dose.

The aims of this study were (1) to establish the first local CT diagnostic reference levels in Kosovo, based on a retrospective audit of a single emergency and ambulatory CT scanner at the University Clinical Centre of Kosovo (UCCK), and (2) to evaluate scan length as an independent dose determinant, comparing per-acquisition CTDIvol and scan length against BfS 2022 reference values and regional comparators.

## Methods

### Study design and setting

This retrospective single-center audit was conducted at the University Clinical Centre of Kosovo, Pristina, which serves as the country’s principal tertiary referral center for both emergency department and ambulatory outpatient imaging. All CT examinations performed between 1 January and 11 March 2026 were eligible for inclusion. The study was approved by the Kosovo Doctors Chamber Ethics Committee (ref. 119/2026, 3 April 2026). Individual informed consent was waived due to the retrospective, fully anonymized nature of the audit, with no alteration to patient care.

### CT scanner

All examinations were performed on a single CT scanner: Philips Incisive CT 128-slice (64 detector rows with z-flying focal spot; software version CHESS5_0; Philips Healthcare, Best, The Netherlands). Single collimation width was 0.625 mm with total collimation widths of 10, 20, or 40 mm depending on protocol. All acquisitions were helical (spiral). Tube voltage ranged from 80 to 140 kV across protocols. CTDIvol was referenced to the 16 cm phantom for head examinations and the 32 cm phantom for body examinations, consistent with IEC 60601-2-44 specifications displayed on the scanner console.

### Data extraction

Dose parameters were extracted directly from DICOM headers using pydicom (v2.4), an open-source Python library for DICOM metadata. The following parameters were extracted per acquisition: CTDIvol, DLP, scan length, tube voltage (kVp), tube current-time product (mAs),single and total collimation width, spiral pitch factor, acquisition type, phantom type, and estimated dose saving (iterative reconstruction). Scan length was derived from the DICOM-reported scan length in millimeters, converted to centimeters. All extracted values were validated against the scanner console dose report display, with 100% concordance. No optical character recognition or manual transcription was used.

### Exclusion criteria

Pediatric patients (age <18 years) were excluded (n = 85). Bolus-tracking monitoring acquisitions were identified by the combination of sequential acquisition type and spiral pitch factor of zero, and were excluded prior to analysis (n = 310). CT-guided biopsy cases (n = 3 acquisitions from 1 patient) were excluded as non-diagnostic. Low-dose chest CT acquisitions (n = 80) were also excluded as this protocol was used for CT-guided biopsy needle placement rather than diagnostic imaging at this institution. Acquisitions recorded on mismatched CTDI phantoms — body phantom (32 cm) acquisitions within head CT studies and head phantom (16 cm) acquisitions within body CT studies, representing additional body regions scanned under a single StudyDescription in polytrauma patients — were identified and excluded (n = 108).

Finally, lumbar spine (n = 23), isolated pelvis (n = 25), head/neck soft tissue (n = 32), and carotid CTA (n = 4) protocols were excluded from DRL analysis due to insufficient sample size or absence of a published comparator. Lumbar spine was specifically excluded because protocol labels did not reliably reflect actual scan extent.

### Examination categorization

Acquisitions were classified into examination categories based on the DICOM StudyDescription field. Brain TRAUMA and Brain TRAUMA+C were combined into Head CT, as both used identical acquisition parameters (BRAIN 2MM protocol, pitch 0.4, scan length approximately 25 cm). Chest examinations were maintained as three distinct categories — chest CT (THORAX NATIVE+C), pulmonary embolism CT, and thorax/abdomen/pelvis CT — to reflect distinct clinical indications with separate BfS comparators. Abdomen CT was renamed abdomen-pelvis CT based on median scan length (57.8 cm), confirming coverage from diaphragm to ischial tuberosities; no pure abdomen protocol was in use at this institution. MSK/Extremity was included in descriptive Table 1 but excluded from Table 3 comparisons due to absence of a BfS comparator and the heterogeneous anatomical regions comprising this category. Nine final examination categories comprising 1,535 acquisitions from 1,092 patients were included in the analysis.

**Table 1.** Per-acquisition dose descriptors and proposed local diagnostic reference levels. CTDIvol (mGy), DLP (mGy·cm), SL = scan length (cm). P75 = 75th percentile (proposed local DRL). CTDIvol referenced to 16 cm phantom for head examinations and 32 cm phantom for body. MSK/Extremity included for completeness. Acquisitions on mismatched phantoms, lumbar spine, and CT-guided biopsy cases excluded (see Methods).

| Exam Category | n | CTDIvol<br>Median | CTDIvol<br>P75 | DLP<br>Median | DLP P75 | SL<br>Median | SL P75 |
| --- | --- | --- | --- | --- | --- | --- | --- |
| Head CT | 590 | 52.4 | 59.4 | 1366.5 | 1701 | 25.8 | 29.6 |
| Chest CT | 187 | 12 | 14.1 | 528.2 | 690.4 | 44.4 | 50.6 |
| Abdomen-Pelvis CT | 369 | 12.1 | 13 | 701.2 | 760.8 | 57.8 | 61.1 |
| Thorax/Abd/Pelvis CT | 61 | 16 | 18.2 | 1167.8 | 1346.2 | 74.4 | 80.3 |
| Pulmonary Embolism CT | 56 | 13.6 | 15.9 | 604.5 | 789.8 | 44.7 | 49.7 |
| Brain CTA | 48 | 42.8 | 58.6 | 1670.5 | 1990.3 | 37 | 43.3 |
| CTA (Aorta) | 92 | 11.2 | 12.8 | 1246.1 | 1541.7 | 130.4 | 141.6 |
| Coronary CTA | 40 | 95.4 | 98.2 | 1990 | 2120.3 | 21.6 | 22.4 |
| MSK/Extremity | 92 | 20.5 | 28.4 | 561.6 | 787.5 | 27.2 | 30.8 |

### Dose descriptors and local DRLs

Per-acquisition dose descriptors reported were CTDIvol (mGy), DLP (mGy·cm), and scan length (cm). Proposed local diagnostic reference levels were defined as the 75th percentile (P75) of each dose descriptor per examination category, consistent with ICRP 135 recommendations. (Vañó et al., 2017) Per-patient total DLP was calculated as the sum of DLP values for all acquisitions within a single patient-examination encounter.

### Statistical analysis

Data are presented as median and P75 values. Per-acquisition CTDIvol and scan length were compared against BfS 2022 reference values using the one-sample Wilcoxon signed-rank test (two-sided), with significance set at p < 0.01. P75 values for CTDIvol and DLP were compared descriptively against Kahraman 2024 local DRLs from Türkiye where available. (Kahraman et al., 2024) Spearman rank correlation was used to assess the association between scan length and DLP. Patient weight data were not available, precluding size-specific dose estimates (SSDE) or weight-stratified analysis. All statistical analyses were performed in R (version 4.5.3; R Foundation for Statistical Computing, Vienna, Austria). Claude (Anthropic, version Claude Opus) was used to assist with statistical analysis scripting and manuscript drafting. All outputs were reviewed, verified, and edited by the authors, who take full responsibility for the content.

### Comparators

Two external reference sets were used. The BfS 2022 diagnostic reference levels (BfS, 2022) provide CTDIvol as the formal DRL quantity and scan length as an orientation value — explicitly designated as guidance rather than a formal DRL per the BfS Leitfaden. (BfS, 2023) The BfS framework defines scan ranges for each examination type; for brain CT, this is skull base to calvarium (13 cm), with no extended trauma protocol. For combined trauma imaging, separate acquisitions with region-specific parameters are recommended. These represent the only published national reference that includes scan length data for CT examinations. Kahraman 2024 local DRLs from Başkent University, Türkiye (104,272 examinations across 8 scanners) (Kahraman et al., 2024) were used as a regional comparator, providing P75 values for CTDIvol and DLP for head, chest, abdomen-pelvis, and coronary CTA examinations. No CT dose data from the Western Balkans were available for comparison.

## Results

After all exclusions, 1,535 acquisitions from 1,092 patients (596 male, 496 female; median age 60 years, IQR 43–71) were analyzed across nine examination categories. Per-acquisition dose descriptors and proposed local DRLs are presented in Table 1. Per-patient total DLP and phase counts are presented in Table 2. Comparison with BfS 2022 and Kahraman 2024 references is presented in Table 3. In-text comparisons use median values to characterize typical practice; Table 3 presents P75-based comparisons for DRL benchmarking.

**Table 2.** Per-patient total DLP and phase count. Total DLP (mGy·cm) = sum of all acquisitions per patient-exam. BfS 2022 does not provide DLP reference values.

| Exam Category | Patients | Mean Acq<br>/Patient | Phases<br>Median | Phases<br>P75 | Total DLP<br>Median | Total DLP<br>P75 |
| --- | --- | --- | --- | --- | --- | --- |
| Head CT | 565 | 1 | 1 | 1 | 1377.8 | 1748.5 |
| Chest CT | 121 | 1.5 | 1 | 2 | 702.9 | 1164 |
| Abdomen-Pelvis CT | 160 | 2.3 | 3 | 3 | 1588 | 2128.9 |
| Thorax/Abd/Pelvis CT | 31 | 2 | 2 | 3 | 2374.3 | 3150.1 |
| Pulmonary Embolism CT | 25 | 2.2 | 2 | 2 | 1363.9 | 1615 |
| Brain CTA | 22 | 2.2 | 2 | 2 | 3993.8 | 5164.5 |
| CTA (Aorta) | 43 | 2.1 | 2 | 2 | 2637.1 | 3150.8 |
| Coronary CTA | 38 | 1.1 | 1 | 1 | 1990 | 2143.3 |
| MSK/Extremity | 87 | 1.1 | 1 | 1 | 578.9 | 817.3 |

**Table 3.** Comparison with BfS 2022 and Kahraman 2024 (Türkiye) references. CTDIvol (mGy) and DLP (mGy·cm) compared as P75 values against Kahraman 2024 local DRLs (P75) where available. SL (cm) compared against BfS 2022 orientation values. — = no comparator available. MSK/Extremity excluded.

| Exam Category | n | CTDIvol<br>P75 | BfS<br>CTDIvol | Kahraman<br>CTDIvol | DLP<br>P75 | Kahraman<br>DLP | SL<br>P75 | BfS<br>Orient. | SL<br>Diff<br>(%) |
| --- | --- | --- | --- | --- | --- | --- | --- | --- | --- |
| Head CT | 590 | 59.4 | 55 | 41.2 | 1701 | 839 | 29.6 | 13 | +127.7 |
| Chest CT | 187 | 14.1 | 8 | 9.3 | 690.4 | 364.8 | 50.6 | 31 | +63.2 |
| Abdomen-Pelvis<br>CT | 369 | 13 | 12 | 11.2 | 760.8 | 588.9 | 61.1 | 45 | +35.8 |
| Thorax/Abd/Pelvis<br>CT | 61 | 18.2 | 12 | — | 1346.2 | — | 80.3 | 63 | +27.5 |
| Pulmonary<br>Embolism CT | 56 | 15.9 | 8 | — | 789.8 | — | 49.7 | 31 | +60.3 |
| Brain CTA | 48 | 58.6 | 15 | — | 1990.3 | — | 43.3 | 33 | +31.2 |
| Coronary CTA | 40 | 98.2 | 20 | 33.4 | 2120.3 | 642.3 | 22.4 | 12 | +86.7 |

### Per-acquisition dose levels

Head CT was the largest category (590 acquisitions, 565 patients). CTDIvol (median 52.4 mGy) was 4.7% below the BfS 2022 DRL of 55 mGy (p = 0.003), while scan length (median 25.8 cm) exceeded the BfS orientation value of 13 cm by 98.5% (p < 0.001). Abdomen-pelvis CTDIvol (median 12.1 mGy) was not significantly different from the BfS reference of 12 mGy (p = 0.95), yet scan length exceeded the reference by 28% (median 57.8 vs 45 cm, p < 0.001).

Coronary CTA demonstrated the highest CTDIvol elevation: median 95.4 mGy versus the BfS DRL of 20 mGy (+377%, p < 0.001). A spiral pitch factor of 0.15 was used in 39 of 40 acquisitions (98%), consistent with retrospective ECG gating. The BfS 2022 framework notes that retrospective gating should be reserved for special circumstances such as arrhythmia, and that any resulting DRL exceedance must be justified. (BfS, 2022) Brain CTA showed the opposite pattern: CTDIvol markedly elevated (+186%, p < 0.001) while scan length was only modestly above reference (+12%, p = 0.003).

All scan length differences were statistically significant (p < 0.01). Chest CT showed CTDIvol +50% and scan length +43% above BfS references. Pulmonary embolism CT showed CTDIvol +70% and scan length +44%. Thorax/abdomen/pelvis CT scan length exceeded the BfS Rumpf orientation by 18%. CTA aorta showed substantial scan length excess (+114%), reflecting inclusion of both isolated aortic and full aorta-to-feet runoff studies within this category.

### Per-patient total dose

Abdomen-pelvis patients received a median of 3 acquisition phases per examination, with median total DLP of 1,588 mGy·cm (P75: 2,129 mGy·cm). Brain CTA patients received the highest per-patient total DLP (median 3,994 mGy·cm across a median of 2 phases). Head CT patients predominantly received a single acquisition (median 1 phase), with median total DLP of 1,378 mGy·cm. Thorax/abdomen/pelvis patients received a median total DLP of 2,374 mGy·cm across 2 phases.

### Scan length as dose driver

Spearman correlation between scan length and DLP was significant across all categories (p < 0.05). The strongest correlations were observed for CTA aorta (ρ = 0.78), head CT (ρ = 0.71), thorax/abdomen/pelvis CT (ρ = 0.60), and abdomen-pelvis (ρ = 0.59).

## Discussion

This study analyzed 1,535 acquisitions from 1,092 patients examined on a single emergency and ambulatory CT scanner at UCCK. These results provide an institutional baseline against which future optimization efforts can be measured.

### Scan length as the primary dose driver

The most notable finding is the systematic excess in scan length across nearly all examination categories, despite CTDIvol values that were, for several protocols, comparable to or below established European references. Head CT exemplifies this dissociation: CTDIvol was 4.7% below the BfS 2022 DRL, yet scan length exceeded the BfS orientation value by 98.5% (median 25.8 cm vs 13 cm) (Figure 1). Since DLP is the product of CTDIvol and scan length, this excess directly translates to patients receiving approximately twice the necessary dose-length product — not because of inappropriate beam intensity, but because the scan extends beyond the anatomical region of interest.

**Figure 1.**
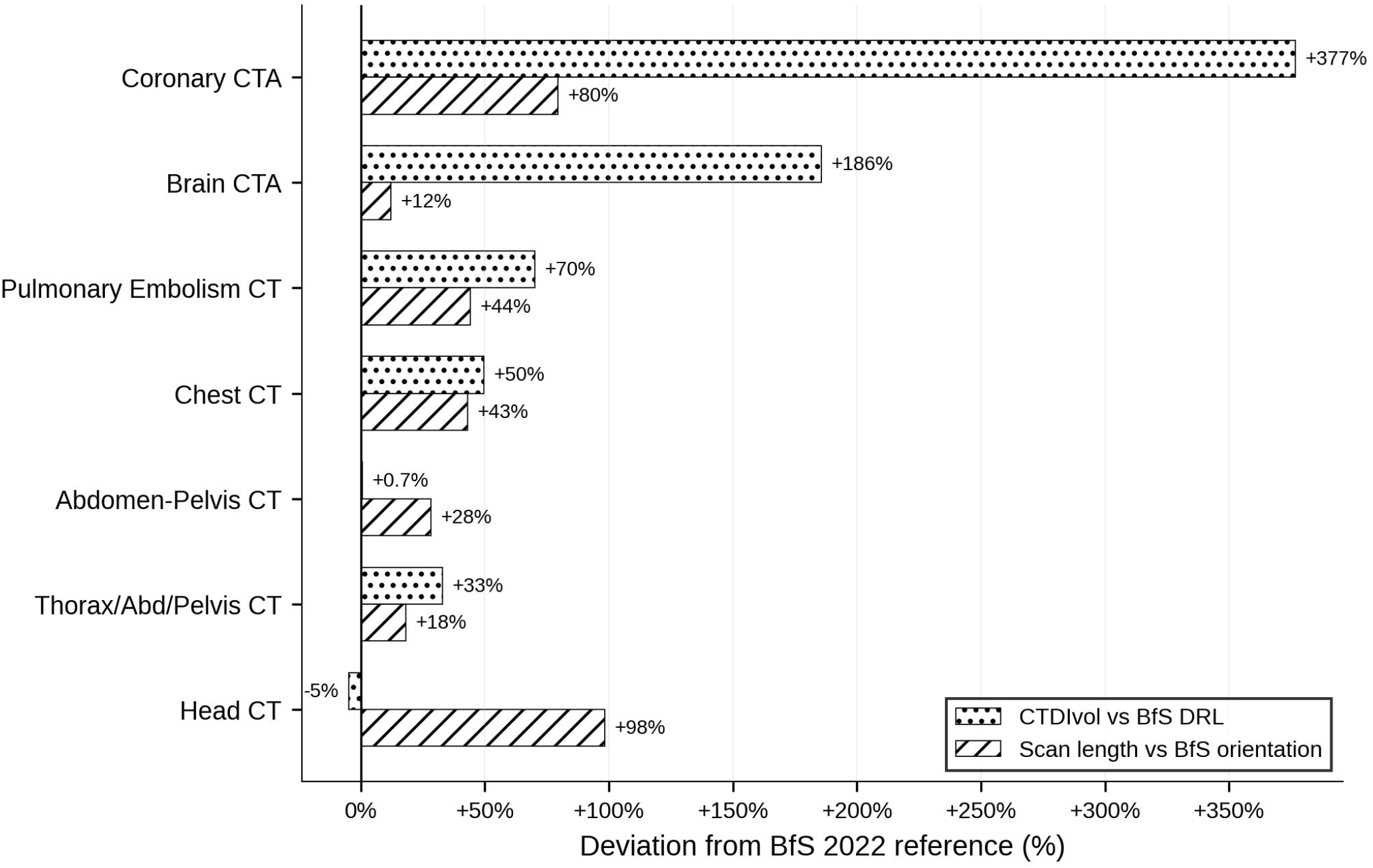
Percentage deviation of median CTDIvol and scan length from BfS 2022 reference values, by examination category. CTDIvol compared against BfS diagnostic reference levels; scan length compared against BfS orientation values. CTA aorta excluded due to mixed study types (see text).

This pattern was consistent across categories. Abdomen-pelvis CTDIvol was essentially identical to the BfS reference (p = 0.95), yet scan length exceeded it by 28%. A CTDIvol-only framework would have classified this protocol as optimized, missing the scan length excess entirely.

The BfS 2022 update is directly relevant. Germany replaced DLP with CTDIvol as the sole DRL metric and introduced scan length as an orientation parameter, explicitly acknowledging for the first time that scan coverage is a critical variable. (BfS, 2022; BfS, 2023) Our findings provide empirical support for this shift.

### Head CT: systematic overscanning across all referral sources

The BfS defines brain CT scan range as skull base to calvarium (13 cm) and does not include a trauma-specific extended head protocol. (BfS, 2022) If cervical spine imaging is clinically indicated, a separate acquisition with body protocol parameters is recommended. The median scan length of 25.8 cm at our institution indicates routine coverage extending from the upper cervical spine through well above the vertex.

Scan length excess was consistent across referral sources: emergency department referrals (74% of patients; median 26.2 cm), neurology (13%; median 24.8 cm), and all other departments showed scan lengths approximately double the BfS orientation value. Excluding emergency referrals, median head CT scan length remained 24.8 cm, indicating that the observed excess is attributable to systematic landmarking practice rather than emergency-specific clinical requirements. This finding is actionable and zero-cost: refining scan start and end landmarks through technologist training requires no equipment upgrade or software modification. Single-site CT dose audits have been shown to provide clinically relevant insights for local optimization, even in the absence of national DRL frameworks. (Tan et al., 2023)

### Coronary CTA: gating technique

The most extreme CTDIvol elevation was observed in coronary CTA (+377%, median 95.4 vs BfS 20 mGy). A spiral pitch factor of 0.15 was used in 98% of acquisitions, consistent with retrospective ECG gating, which irradiates continuously throughout the cardiac cycle. (Hausleiter et al., 2009) The BfS explicitly states that retrospective gating should only be performed under special circumstances (e.g., patients with arrhythmia) and that resulting DRL exceedances must be justified. (BfS, 2022) The P75 CTDIvol of 98.2 mGy approaches the BfS action threshold for reporting significant events. Transition to prospective ECG triggering, where clinically appropriate, would substantially reduce dose. The DLP P75 of 2,120 mGy·cm compared with Kahraman’s 642 mGy·cm (Kahraman et al., 2024) (3.3-fold difference) reinforces this as a priority for protocol review. However, in the absence of heart rate or arrhythmia data, the clinical necessity of retrospective gating in individual cases cannot be assessed.

### Iterative reconstruction

Iterative reconstruction was not enabled for head CT, brain CTA, coronary CTA, or MSK protocols (0% dose saving in 100% of acquisitions) despite being active for body protocols (27– 46% saving). Enabling iterative reconstruction would permit reduction of tube current while maintaining diagnostic image noise, directly lowering CTDIvol.

### Abdomen-pelvis: optimized CTDIvol, excess scan length

Abdomen-pelvis CTDIvol matched the BfS reference, with 100 kV used in 97% of acquisitions. The 28% scan length excess indicates room for landmarking refinement in this otherwise well-optimized category.

### Multi-phase imaging and total patient dose

Per-patient total DLP reflected multi-phase imaging practice (Table 2), with brain CTA patients receiving the highest cumulative exposure (median 3,994 mGy·cm across 2 phases). BfS 2022 intentionally omitted DLP as a DRL metric. (BfS, 2022)

### Comparison with regional data

Our proposed DRLs can be compared with the Kahraman 2024 local DRLs from Türkiye, the geographically closest available comparator. (Kahraman et al., 2024) Head CT CTDIvol P75 (59.4 mGy) exceeded the Kahraman value (41.2 mGy) by 44%. Abdomen-pelvis CTDIvol P75 (13.0 mGy) was comparable to Kahraman (11.2 mGy). Coronary CTA showed the largest discrepancy: P75 of 98.2 mGy versus Kahraman’s 33.4 mGy, consistent with the gating technique difference discussed above. Kahraman did not report scan length data, precluding direct comparison on this metric — a gap that highlights why the BfS approach of reporting scan length as a complementary metric is needed.

## Limitations

This study has several limitations. First, data were collected from a single CT scanner at one institution, limiting generalizability to a national DRL. However, as the first CT dose data from Kosovo, these local DRLs serve as a necessary starting point. Second, patient weight was not routinely recorded in the DICOM header, precluding size-specific dose estimates and weight-stratified analysis. The scanner calculates and displays SSDE from the surview-derived effective diameter; however, SSDE is derived from a single-dimension size estimate rather than actual patient weight. (AAPM, 2011) Weight-stratified analysis and validated SSDE reporting should be incorporated into future prospective audits through mandatory weight entry in the scanner’s patient registration workflow. Third, certain categories had limited sample sizes (brain CTA n = 48, coronary CTA n = 40), which should be considered when interpreting the proposed DRLs. Fourth, CTA aorta combines isolated aortic with full runoff studies, inflating scan length variability. Fifth, BfS scan length values are orientation parameters, not formal DRLs; however, they represent the only published national reference for this metric. Sixth, no image quality assessment was performed; dose optimization without concurrent image quality evaluation is incomplete. However, no clinical concerns regarding diagnostic adequacy have been raised at this institution, suggesting that dose reduction through scan length optimization may be achievable without compromising diagnostic performance. A prospective audit pairing scan length reduction with systematic image quality evaluation is planned. Seventh, the retrospective design precludes assessment of clinical justification for individual examinations. Eighth, generic StudyDescription fields precluded indication-based DRL analysis as recommended by international frameworks. (Bos et al., 2022) Finally, the 10-week study period may not capture seasonal or referral pattern variation, though the sample size is sufficient for P75 estimation per ICRP recommendations. (Vañó et al., 2017)

## Conclusion

This first CT dose audit from Kosovo reveals that excess scan length, not CTDIvol, is a major source of elevated patient dose in most examination categories. Head CT scan lengths were approximately twice the BfS orientation value regardless of referral source, identifying systematic landmarking practice as the primary target for optimization. Coronary CTA doses were markedly elevated due to routine retrospective ECG gating. Iterative reconstruction was not enabled for head or cardiac protocols despite being available on the scanner.

These findings are correctable through technologist landmarking training, protocol review for coronary CTA gating technique, and enabling iterative reconstruction — interventions requiring no equipment investment. The proposed local DRLs and scan length data provide a pre-intervention baseline for a structured optimization program at UCCK. Extension to additional CT scanners within Kosovo would enable establishment of national DRLs. These data may also serve as an initial reference for other Western Balkan institutions seeking to establish their own dose benchmarks.

## Data Availability

All data produced in the present work are contained in the manuscript

## Acknowledgements

None.

## Funding

This research did not receive any specific funding.

## Conflicts of interest

The authors declare that they have no conflict of interest.

## Data availability statement

The research data associated with this article are included within the article.

## Author contribution statement

G. Rudi: Conceptualization, Data extraction, Data analysis, Writing — original draft. F. Vula: Data validation, Writing — review and editing. A. Bicaku: Clinical protocol verification, Writing — review and editing. K. Dedushi Hoti: Supervision, Writing — review and editing. I. Ahmetgjekaj: Supervision, Writing — review and editing.

## Ethics approval

This study was approved by the Kosovo Doctors Chamber Ethics Committee (ref. 119/2026, 3 April 2026).

## Informed consent

Individual informed consent was waived due to the retrospective, fully anonymized nature of the audit, with no alteration to patient care.

## Notes

### Competing Interest Statement

The authors have declared no competing interest.

### Funding Statement

This study did not receive any funding

### Author Declarations

The study was approved by the Kosovo Doctors Chamber Ethics Committee (ref. 119/2026, 3 April 2026).

